# Pharmacogenomic Drug-Gene Interactions in Geriatric Emergency Department Patients That Have Fallen

**DOI:** 10.1101/2024.08.14.24311970

**Authors:** Richard D. Shih, Gabriella Engstrom, Abhijit S. Pandya, Gregg B. Fields, Borivoje Furht, Ali A. Danesh, Scott M. Alter, Humberto Munoz BPharm, Lisa M. Clayton, Joshua J. Solano, Timothy Buckley, Olivia Hung, Alexander Farag, Mike Wells

## Abstract

**Background:** Pharmacogenomic assisted prescribing of medications utilizes individual genetic information to identify drug-gene interactions. We aimed to assess potential pharmacogenomic drug-gene interactions in geriatric emergency department (ED) patients that have sustained a fall.

**Methods:** This was a prospective study involving 25 older adult ED patients with fall related injury. Data collected included current medications, demographics and mechanism of injury. All patients provided a DNA sample for pharmacogenomic testing, MatchMyMeds (DNA Labs, Boca Raton, FL) which assessed genetic data for 23 enzyme systems and reports on potential drug-gene interactions for 134 medications. Each patients’ medications were reviewed against their pharmacogenomic report and categorized as Green (go), Yellow (caution) or Red (stop) based on their genetic information and published interactions by the Clinical Pharmacogenetics Implementation Consortium (CPIC), Dutch Pharmacogenetics Working Group (DPWG) and Food and Drug Administration-approved drug label information. The main study outcome was pharmacogenomic drug-gene interactions.

**Results:** Of the 25 patients enrolled (median age, 81 years, IQR: 76-85), 68% were female. Patients were taking a median of 8 medications (IQR: 5-11). The most common types were antihypertensives, statins, anticoagulants, and anti-platelet medications. Significant drug-gene interactions (Yellow or Red) were identified in 14/25 (56%) patients. Further, 6/25 (24%) had one or more potentially serious (Red) interactions identified.

**Conclusions:** In geriatric ED patients with a fall-related injury, a majority have a significant pharmacogenomic drug-gene interactions. DNA testing identifies these interactions and can assist with pharmacogenomic-guided medication prescribing which may decrease ADEs and improve clinical outcomes.

**Key Points:**

- Polypharmacy is associated with adverse drug events and fall-related injuries.
- Pharmacogenomic can identify drug-gene interactions in older ED adults that sustain a fall.
- Significant drug-gene interactions are common in older ED adults that sustain a fall.
- Pharmacogenomic-guided medication prescribing which may decrease ADEs and improve clinical outcomes

## Introduction

As individuals age, they have more medical problems and are prescribed more medications. Taking more than five medications, i.e., polypharmacy, leads to increased adverse drug events (ADEs), hospitalizations and poor outcomes [1–4]. Of these poor outcomes, falling and fall-related injury are among the most important [1,5].

Stopping a medication, i.e., deprescribing, is a very difficult process [6]. Although several methods for medication assessment and possible discontinuation have been published, most have not been widely adopted and there are many challenges associated with stopping medications [7–11]. Pharmacogenomic tests may provide an objective method for physicians to identify medications that are high risk for ADE and allow them to deprescribe or prescribe medications more precisely and appropriately. The objective of this study was to assess pharmacogenomic drug-gene interactions in geriatric ED patients that have fallen. This may provide the basis for pharmacogenomic-guided medication prescribing in older patients at risk for fall related injury.

## Methods

### Study Design and Setting

We conducted a prospective study involving 25 patients presenting to a single ED with an annual patient volume of 50,000. The study was approved by the hospital’s affiliated university institutional review board.

### Selection of Participants

A convenience sample of ED patients aged 65 years or older who had sustained a fall related injury were eligible and consented for participation. We excluded patients with penetrating injuries, injury more than 24 hours prior to presentation and transfer from another hospital. Additionally, we excluded any patients that were hospice and/or Do Not Resuscitate (DNR) status.

### Data Collection

Identified patients who met the study’s inclusion and exclusion criteria were approached to complete an IRB-approved informed consent form for study participation. Data were collected by trained research assistants (RAs), who were present in the ED. Data collected included current medications (prescribed pre-injury), demographic data, mechanism of injury, past medical history, and ED diagnosis.

In addition, all patients provided a DNA sample obtained via a single cheek swab sample from the inside of the patient’s mouth. The subjects were not allowed to eat, drink, or smoke for 30 minutes prior to swabbing. The pharmacogenomic test utilized was MatchMyMeds (DNA Labs, Boca Raton, FL, https://dnalabs.ca). This test analyzes for 23 different metabolic enzyme systems and reviews potential drug-gene interactions for 134 different medications (see Table 1). The MatchMyMeds report is based on the Clinical Pharmacogenetics Implementation Consortium (CPIC, https://cpicpgx.org) [12], the Dutch Pharmacogenetics Working Group (DPWG, https://www.pharmgkb.org/page/dpwg) [13,14], and Food and Drug Administration-approved drug label information to provide detailed drug and dosing recommendations. In addition, based on the individual’s genetics, the report categorized 134 medications as Green (go: use label recommended dosage and administration), Yellow (caution: use with caution - read detailed recommendations for potential dose adjustment) or Red (stop: select alternative treatment if possible - read detailed recommendation for detail).

**Table 1.**
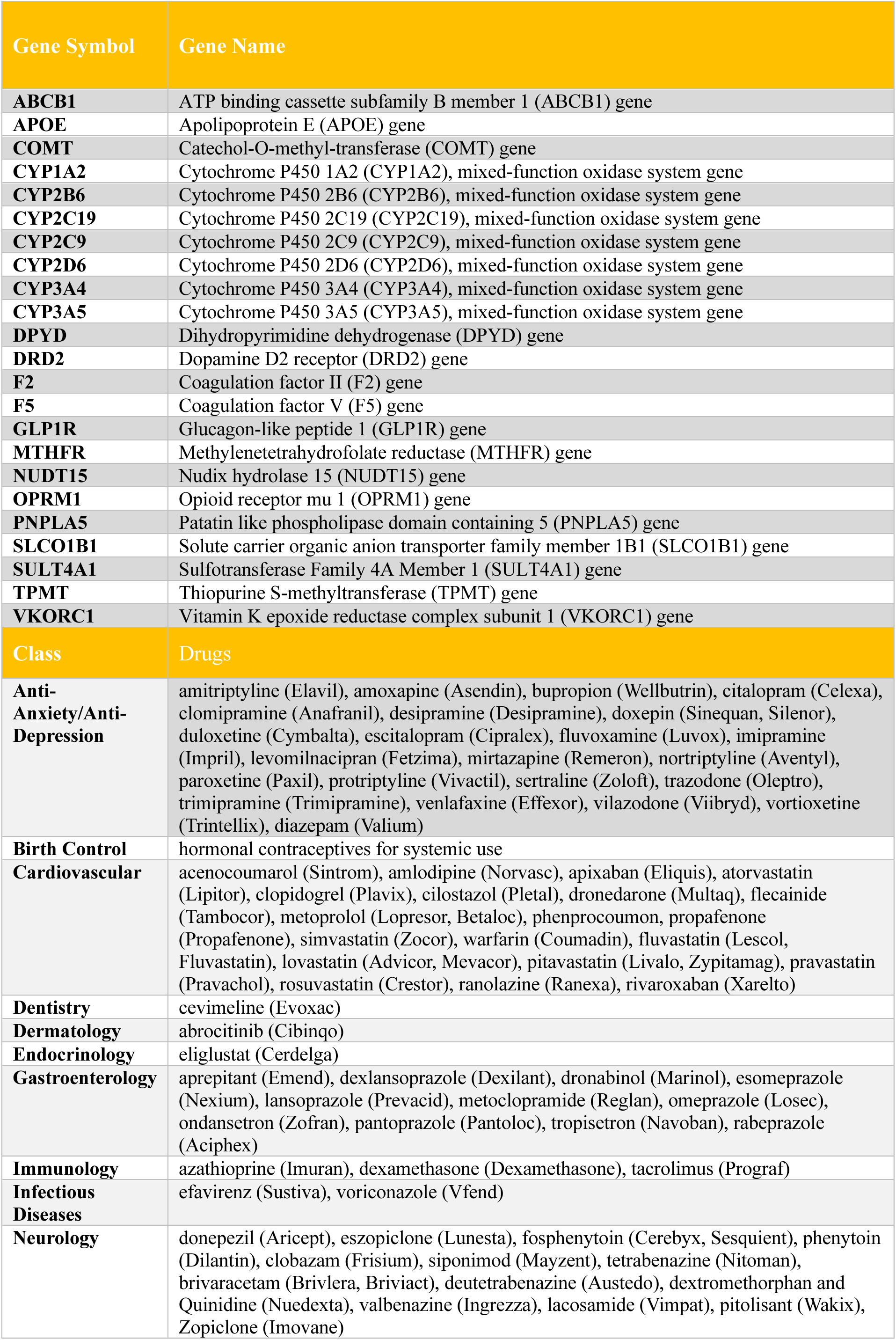
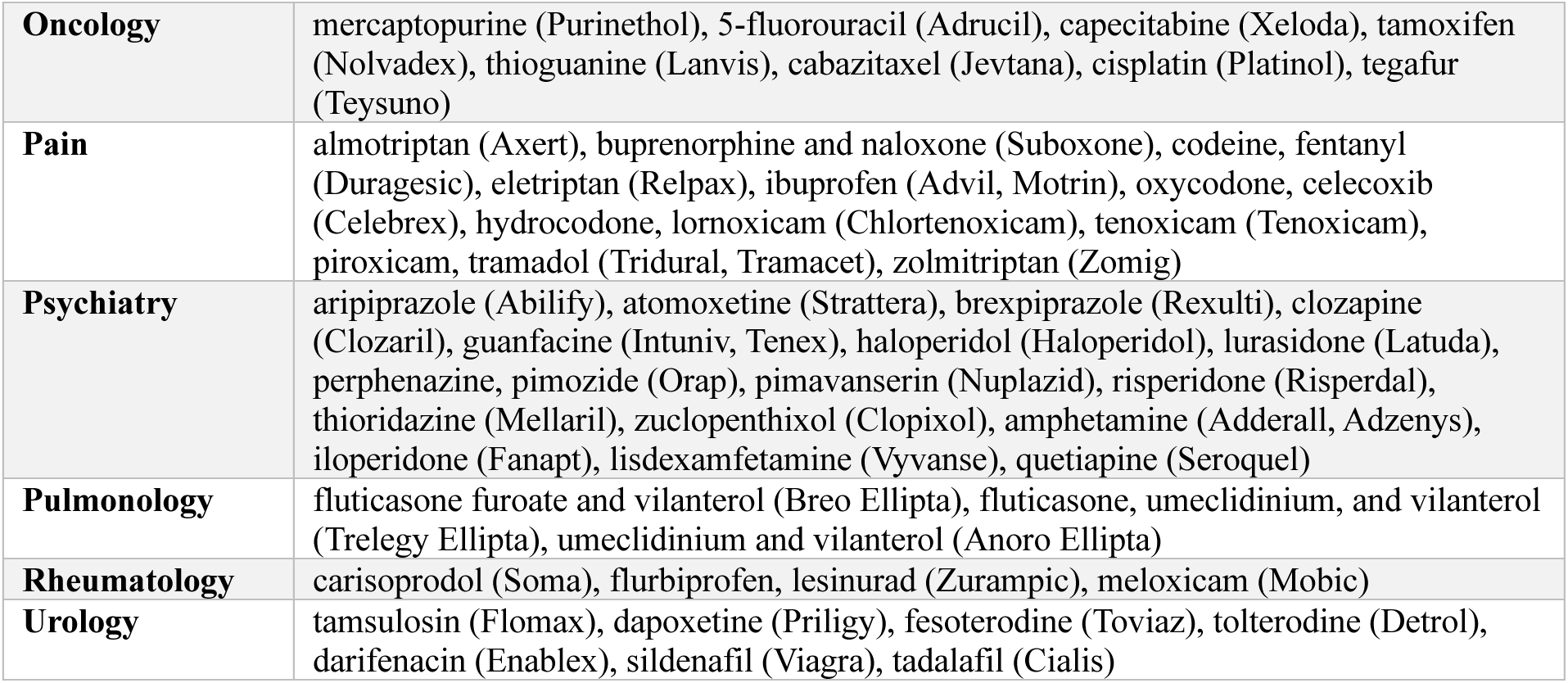
Metabolic Enzyme System Genes Analyzed and Medications Assessed for Drug-Gene Interactions.

### Outcomes

The main study outcome was to identify pharmacogenomic drug-gene Interactions. Yellow and Red medications were defined as serious interactions.

A secondary outcome was to assess whether a medication with a significant gene-drug interaction was also identified on the Beers potentially inappropriate medications list [15].

### Analysis

Descriptive analyses were conducted to assess the prevalence and severity of drug-gene interactions, as well as to identify the specific drugs affected. The frequency of interactions within each risk category (Green, Yellow, and Red) was quantified, and the severity of these interactions was characterized based on their potential impact on patient safety and treatment efficacy.

## Results

Twenty-five patients were enrolled. The median age was 81 years (IQR: 76-85) and 68% were female. The most frequent co-morbidities were hypertension (64%), atrial fibrillation (32%), coronary artery disease (28%) and stroke (8%).

All patients were on medications for pre-existing co-morbidities (see Table 2). Patients were taking a median of 8 medications (IQR: 5-11). The most common types were antihypertensives, statins, anticoagulants, and anti-platelet medications. All the patients had suffered a fall, as this was part of the inclusion criteria. Most had contusions, fractures and wounds. Several also had significant medical problems such as stroke, head injury and syncope (see Table 3).

**Table 2:** Clinical Characteristics.

| Patients |  |
| --- | --- |
| N=25 |  |
| Past Medical History, n (%) |  |
| Hypertension | 16 (64) |
| Atrial fibrillation | 8 (32) |
| Coronary artery disease | 7 (28) |
| Solid tumor/Cancer active | 4 (16) |
| Cerebrovascular attack | 2 (8) |
| Valve replacement | 2 (8) |
| Myocardial infarction | 2 (8) |
| Diabetes | 1 (4) |
| Chronic obstructive pulmonary disease | 1 (4) |
| Congestive heart failure | 1 (4) |
| Pulmonary embolism | 1 (4) |
| Peripheral vascular disease/peripheral stent | 1 (4) |
| Chronic kidney disease | 1 (4) |

**Table 3:**
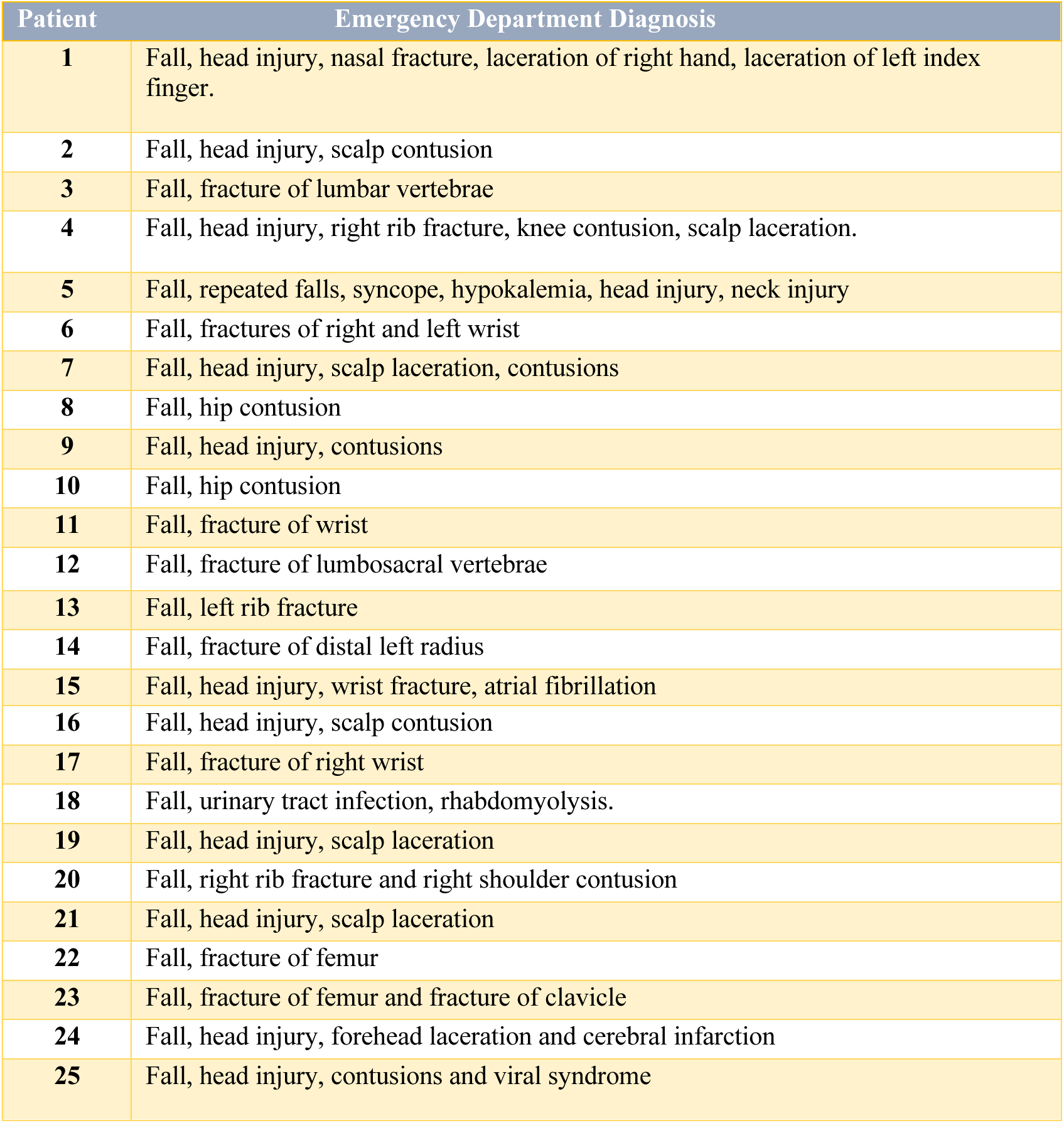
Patients’ Emergency Department Diagnosis.

When considering the pharmacogenomic data and drug-gene interactions, of our 25 patients, 14/25 (56%) had one or more Yellow or Red medications (see figure 1) for a total of 20 significant gene-drug interactions (see Table 4). Further, 6/25 (24%) had one or more Red medications identified (see Figure 1). The implicated medications were those that could increase risk of falling with several drugs with the potential for increased sedation (escitalopram, tramadol, trazodone, oxycodone and duloxetine) and others potentially causing bradycardia or hypotension (metoprolol and amlodipine).

**Figure 1.**
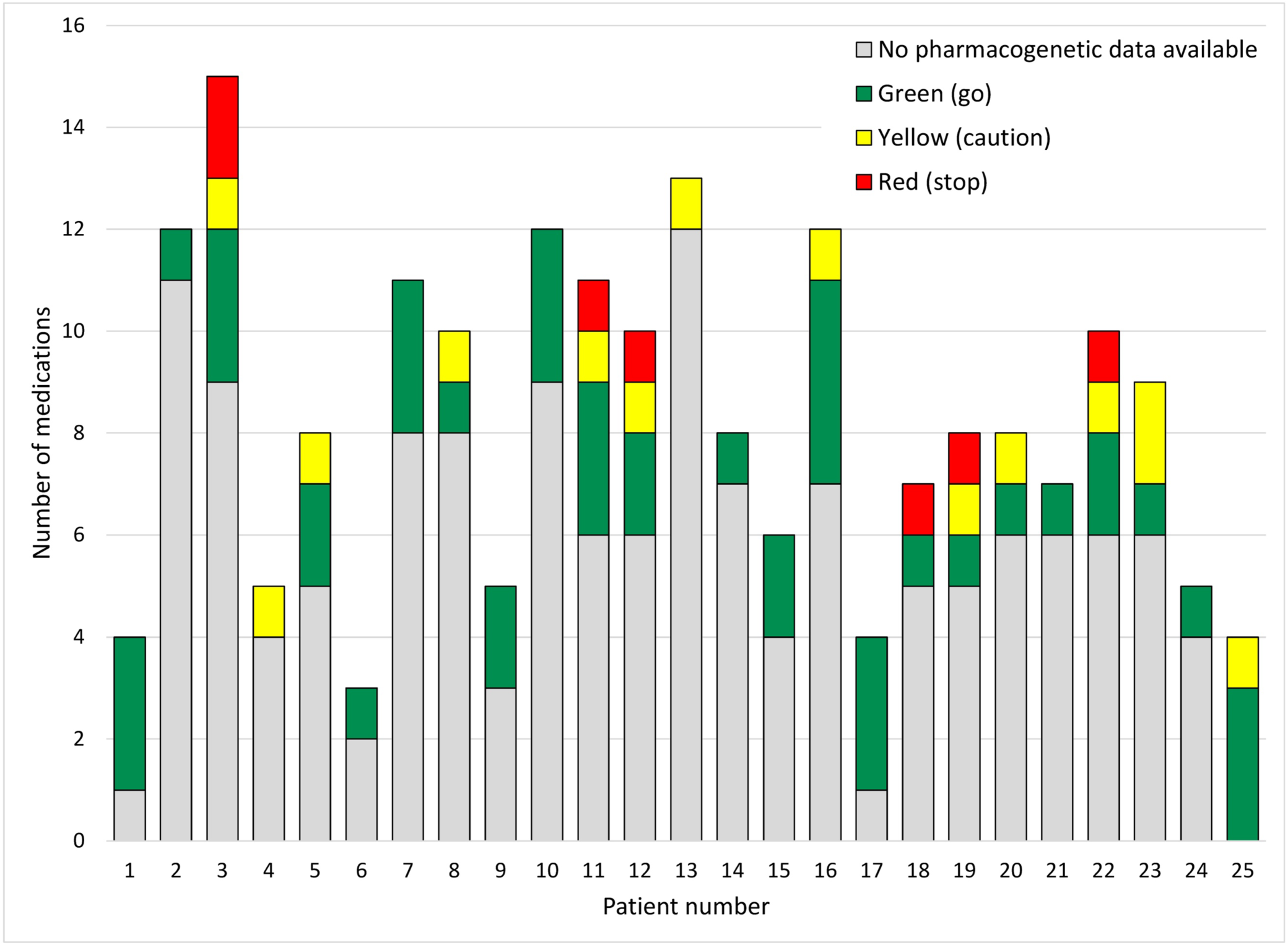
Possible drug-gene medication interactions for each patient.

**Table 4:**
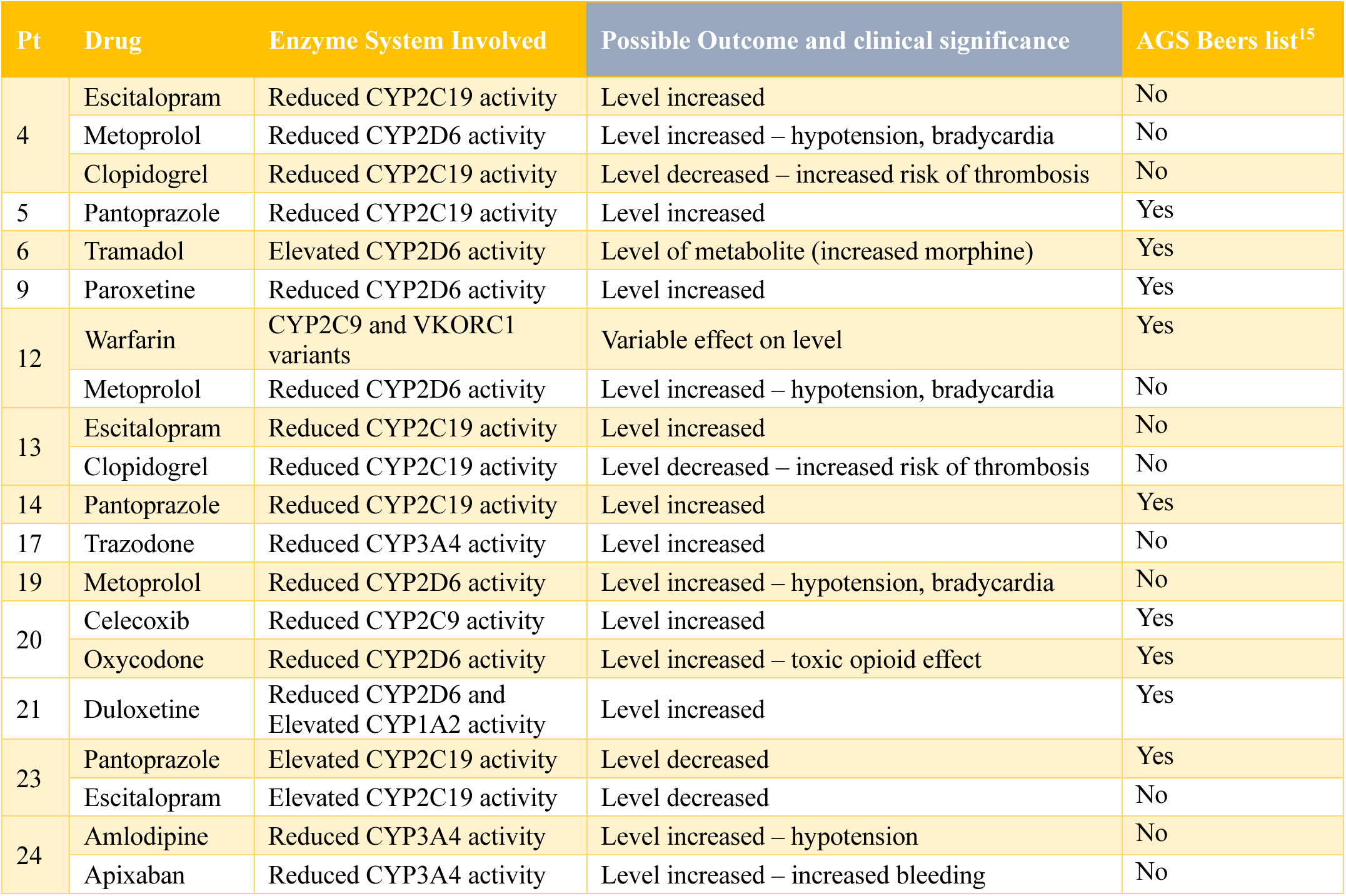
Significant Drug-Gene Interactions.

## Discussion

As an individual gets older, they develop more medical problems and are prescribed more medicines. The more medications that someone takes, the higher the risk for an ADE [16]. Polypharmacy, taking five or more medicines is a major risk factor for an ADE [17].

Shaver et. al. in 2021 showed that mortality risk in older United States (US) persons from falls increased steadily from 1999 to 2017, as the use of Fall Risk Increasing Drugs (FRIDs) increased. During that period, the percentage of older people taking a FRIDs increased from 57% to 97% [3].

On average, 750 US older adults are hospitalized each day due to an ADE and 50% are taking seven or more medications [5,11]. Further, about 60% of older people have one or more medicines that are unnecessary [18]. Among geriatric ED patients about 75% have one or more medications that are classified as “avoid” or “use with caution [6].”

However, stopping a medication (deprescribing) has proven to be very difficult for patients and physicians [11]. Although a number of methods for medication assessment and possible discontinuation have been published, most have not been widely adopted [7–9,15,19]. Deprescribing presents several significant challenges in clinical practice including doctors lacking training in stopping medications, doctors worrying about worsening symptoms, patients worrying about worsening symptoms, concern for withdrawal, the time required to discuss medication changes, and patients feeling like their doctor is “giving up” and no longer treating their disease [11,12,20].

Pharmacogenomics may provide an objective means to help with this problem. It is a genetic means to assess for possible risk of ADEs and may one day become standard information for physicians to help provide more precise and personalized prescribing of medications. Pharmacogenomics involves gene testing to assess how an individual metabolizes medications. Hepatic enzymes in the P450 systems (i.e. CYPD6, CYP3A4, CYP2C19) are responsible for metabolizing many commonly used medications (i.e. Lipitor®, Valium®, warfarin, Xarelto®, Zoloft®, and many others). In addition, CYP2D6 has many different genetic polymorphisms and depending on the inherited alleles the individual CYP2D6 system may function as poor, intermediate, extensive or ultrarapid metabolizers [21].

Most people are intermediate and extensive metabolizers and in our current system of dosing medications, a “one size fits all” approach, the standard dose will provide concentrations in the therapeutic window predictably. However, if they are a poor metabolizer or ultrarapid metabolizer, a given medication may be in the toxic or ineffective concentration range.

In our study each patient’s medications was assessed for a potential for a drug-gene interaction based on DNA Labs pharmacogenomic report. Of our 25 patients, there were a total of 20 (Red or Yellow) significant potential drug-gene interactions identified with 56% having one or more Yellow or Red medications (see Figure and 5). In addition, 20% had at least one red drug-gene medication interaction identified. Examples of significant gene-drug interactions include:

- Reduced CYP2D6 enzyme activity in someone taking metoprolol. These individuals are poor metabolizers of metoprolol and have a three-to-ten-fold higher concentration compared to an extensive metabolizer. Further, poor metabolizers had a five times higher rate of ADE compared to non-poor metabolizers. These ADE include symptomatic bradycardia and dizziness [22].
- Reduced CYP2C19 enzyme activity in someone taking clopidogrel. Approximately 30% of individuals taking clopidogrel have reduced or zero CYP2C19 activity which leads to decreased platelet inhibition effect and worse cardiovascular outcomes [23].

An additional important finding is that many of the gene-drug interactions that were identified are not on the American Geriatrics Society 2023 updated AGS Beers Criteria® (see Table 4) [15]. The medication involved in 11 of the 20 (55%) significant potential drug-gene interactions are not on this Beers list. The Beers Criteria® is a list of potentially inappropriate medications for older adults. It is developed by a panel of interprofessional experts who review the published literature to establish this list. This list does not appear to reflect potential drug-gene medication interactions, as more than 50% of the medicines involved in interactions identified in our study were not on the Beers list.

### Limitations

This study was a small convenience sample of 25 patients without a comparison group. A larger study with more participants would give a more precise percentage of gene-drug interactions and identify more infrequently occurring ones.

A second limitation is that the gene-drug interactions identified are potential ones and ADEs were not examined as part of this study. A randomized controlled trial utilizing the pharmacogenomic assisted prescribing of medications would be the ideal method to assess for ADEs and the clinical impact of the significant gene-drug interactions that were identified.

A third limitation is that even if individual pharmacogenomic data were available, it is not clear if physicians would utilize this for improved prescribing. One of the barriers is the difficulty in interpretating the pharmacogenomic data leading to an increased amount of time and lack of user-friendly guideline information [12,20,24]. This study did not assess this and is another future area of research for the implementation of pharmacogenomic assisted prescribing practices.

## Conclusions

In geriatric ED patients that present for a fall-related injury, a majority have a significant pharmacogenomic drug-gene interactions. DNA testing can identify these interactions and can assist with pharmacogenomic-guided medication prescribing, which has the potential to decrease ADEs and improve clinical outcomes.

## Data Availability

ll data produced in the present study are available upon reasonable written request to the authors.

